# Predicting Risk of Alzheimer’s Diseases and Related Dementias with AI Foundation Model on Electronic Health Records

**DOI:** 10.1101/2024.04.26.24306180

**Authors:** Weicheng Zhu, Huanze Tang, Hao Zhang, Haresh Rengaraj Rajamohan, Shih-Lun Huang, Xinyue Ma, Ankush Chaudhari, Divyam Madaan, Elaf Almahmoud, Sumit Chopra, John A. Dodson, Abraham A. Brody, Arjun V. Masurkar, Narges Razavian

## Abstract

Early identification of Alzheimer’s disease (AD) and AD-related dementias (ADRD) has high clinical significance, both because of the potential to slow decline through initiating FDA-approved therapies and managing modifiable risk factors, and to help persons living with dementia and their families to plan before cognitive loss makes doing so challenging. However, substantial racial and ethnic disparities in early diagnosis currently lead to additional inequities in care, urging accurate and inclusive risk assessment programs. In this study, we trained an artificial intelligence foundation model to represent the electronic health records (EHR) data with a vast cohort of 1.2 million patients within a large health system. Building upon this foundation EHR model, we developed a predictive Transformer model, named *TRADE*, capable of identifying risks for AD/ADRD and mild cognitive impairment (MCI), by analyzing the past sequential visit records. Amongst individuals 65 and older, our model was able to generate risk predictions for various future timeframes. On the held-out validation set, our model achieved an area under the receiver operating characteristic (AUROC) of 0.772 (95% CI: 0.770, 0.773) for identifying the AD/ADRD/MCI risks in 1 year, and AUROC of 0.735 (95% CI: 0.734, 0.736) in 5 years. The positive predictive values (PPV) in 5 years among individuals with top 1% and 5% highest estimated risks were 39.2% and 27.8%, respectively. These results demonstrate significant improvements upon the current EHR-based AD/ADRD/MCI risk assessment models, paving the way for better prognosis and management of AD/ADRD/MCI at scale.

## Introduction

Alzheimer’s disease (AD) and AD-related dementias (ADRD) are irreversible conditions affecting over 6 million people in the United States^1,2^. While the pathogenesis of AD/ADRD is complex, several modifiable cardiovascular risk factors, such as hypertension, smoking, obesity, and diabetes, are recognized to contribute to its pathogenesis and progression^3–6^. Early diagnosis of AD/ADRD or mild cognitive impairment (MCI) are important for a number of reasons. First, the diagnosis may serve as a motivating factor for addressing modifiable risk factors amongst both clinicians and persons living with dementia (PLWD) or mild cognitive impairment (MCI) as it is one of the only currently known ways to slow cognitive decline. Second, when diagnosed earlier, it is easier for advance care planning to occur with input from the PLWD or MCI, and allows for greater planning for the eventual decline, including caregiving situation, housing or moving closer to family, financial planning. While controversial in nature, early diagnosis is also important as the sole disease-modifying FDA-approved therapy currently on the market for AD/ADRD/MCI is only available and shown some efficacy in PLWD or MCI at the early stage of impairment^7^. Accordingly, early identification of AD/ADRD at the prodromal stage is a critical component in optimal care delivery. However, at the same time, there are significant inequities in care, including lower rates of early diagnosis in racial and ethnic minoritized groups and worse management of modifiable cardiovascular disease factors. These inequities are multifactorial, and new methods are needed for systematically improving the healthcare system.

Algorithms and risk models for detecting probable undiagnosed MCI or AD/ADRD are one way to facilitate such early diagnosis and intervention. Approaches with wearable signals^8^, neuroimaging^9^ and blood biomarkers^10^ are promising but cannot be readily applied to the general population due to the cost and effort, and limited use, especially in underserved and historically marginalized communities. Assessing risks with Electronic Health Records (EHR)-based models has the potential for direct integration into EHR systems and clinical workflow at the point of care for every patient. A few retrospectively validated EHR-based models exist to date. eRADAR, a statistical model^11^, which was developed and validated using participants of Adult Changes in Thought study, is the most robust model tested to date in scientific studies, and is undergoing prospective validation and randomized trials^12,13^. One limitation of the eRADAR model however is its focus on using known risk factors including aging, vascular diseases and diabetes, while other potential factors that can improve precision, such as specific medications, lab values, and additional diagnoses are not included. This is particularly important as many of the eRADAR risk factors have correlations with socioeconomic status and racial and ethnic minoritized groups which can reduce sensitivity in these populations. To address these concerns, we developed and validated an EHR-based risk assessment model utilizing the additional information available in the EHR, exhibiting improved performances on both overall population and subgroups with different characteristics.

Recent advancements in deep learning offered neural networks with a stronger capacity to understand the EHR^14–17^. A graph neural network, taking the connections among various EHR features into account, outperformed traditional statistical models and multiple layer perceptron networks in predicting potential AD^18^. However, this study only used cross-sectional EHR without temporal information. Transformer^19^, a powerful network architecture used to represent high-dimensional data like images and languages, is now widely used as the state-of-the-art method to represent sequential relationship across visits and interconnection of information within a single visit^20–22^. The increment in the scale of the Transformer model usually leads to better performance, while it also requires larger training data to achieve generalizable performance^23^. To address the insufficiency of labeled data, a self-supervised learning approach, masked token modeling, can be used to pretrain these large models on a broader domain with unlabeled data^24,25^. Then the large pretrained model, termed *Foundation Model* can be partially or fully finetuned for some downstream prediction tasks with smaller labeled datasets^26,27^. In this study, we employed this framework to develop a **T**ranformer model for predicting **R**isks of **AD**/ADRD/MCI based on **E**HR (**TRADE**), where we pretrained a foundation model with a large-scale inclusive EHR dataset, and finetuned the model with a selective cohort for predicting AD/ADRD/MCI. TRADE outperformed the vanilla Transformer model trained from scratch. Compared to Transformers for EHR in previous literature (e.g. BEHRT^20^ and MED-BERT^21^), TRADE and its base foundation model further incorporated the medications and discretized lab values, in addition to diagnosis codes, to enrich the EHR representation. Also, it was trained with a curated cohort with more medical concepts, visits and longer span of presence in the healthcare system.

Our approach achieved an area under the receiver operating characteristic (AUROC) at 0.772 (95% CI: 0.770, 0.773) in identifying AD/ADRD/MCI patients in 1 year, and 0.735 (95% CI: 0.734, 0.736) in 5 years, specifically among patients aged 65 and above. The positive predictive values (PPV) in 5 years among patients with the top 1% and 5% highest estimated risks were 39.2% and 27.8%. We built a comparison model on the same cohort with eRADAR that has been deployed in clinical practices^11^. We demonstrated that our proposed deep-learning model was more accurate on the overall heldout validation set and subcohorts separated by different characteristics. These results suggest that incorporating our AI foundation model in the EHR system is promising to advance the current identification and screening of patients at high risk for AD/ADRD/MCI.

## Results

### Study participants

We used Electronic Health Record (EHR) data from a large health system, covering 1,288,333 patients with 587 million medical concept tokens from Jan 2013 to Jan 2023 with at least 5 visits. The EHR of each patient includes a sequence of encounters consisting of International Classification of Diseases (ICD-10) diagnostic codes (all diagnoses types, including encounter, problem list, billing and medical history), lab values coded under Logical Observation Identifiers Names and Codes (LOINC) and medications at the therapeutic level (Figure 1a). The onset of AD/ADRD and MCI was defined as the visit with the first occurrences of AD/ADRD/MCI-related diagnosis codes or medication (Table S1).

**Figure 1.**
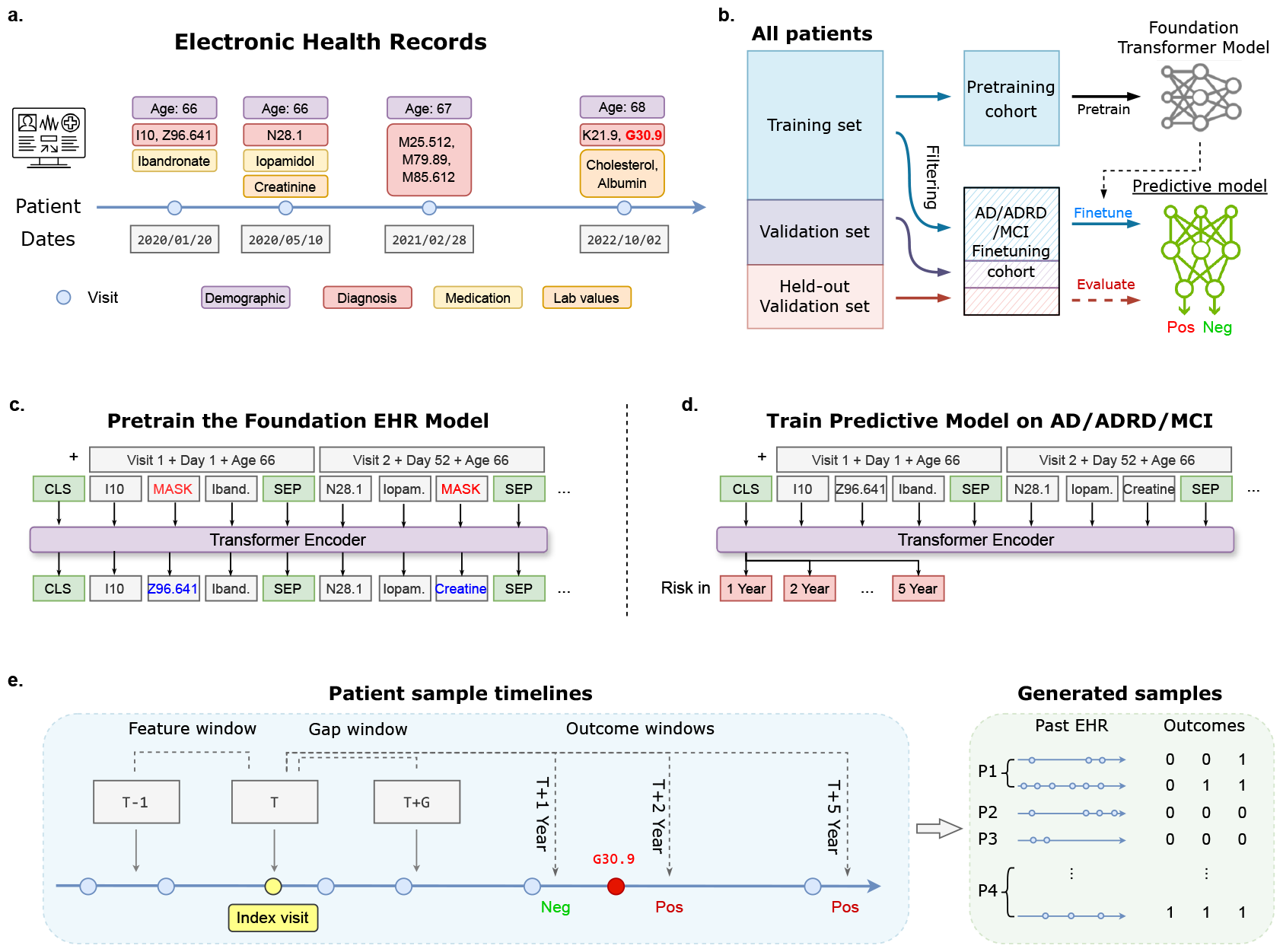
The design of ML architecture of our study. **(a)** The patient’s EHR data of multiple visits, including demographics, diagnoses, lab values, and medication. **(b)** The patients in the study were partitioned into a training, validation, and held-out validation set. The pretraining cohort, formed by patients in the training set, was used to pretrain the foundation Transformer model; the AD/ADRD/MCI finetuning cohort, filtered by age, was used to train and validate the AD/ADRD/MCI predictive model. **(c)** The Transformer model processes variables in EHR as tokens, and encodes indices of visits, days, and ages for 3 distinct positional encodings, which are added to the token embeddings. The foundation model was pretrained by masked token modeling, which taught the model to reconstruct the randomly masked information in the input EHR sequence. **(d)** The AD/ADRD/MCI predictive model was constructed by stacking a linear network upon the output representation of <CLS> token, estimating the risks based on previous EHR. **(e)** The samples for the AD/ADRD/MCI predictive model were generated by sliding windows on various index visits over patient’s longitudinal EHR. We used records in the 1-year feature window prior to the index visit as inputs and the dementia status in various outcome windows following the index visit as labels. Samples with no follow-ups by the end of the outcome window were considered censored and excluded. In some experiments, we specified a minimum gap period between the end of the feature window, and the beginning of the outcome window, and excluded samples with AD/ADRD/MCI onset in the gap window to avoid data leakage due to lengthy AD/ADRD/MCI diagnosis processes.

As shown in Figure 1a, the patients in the study were randomly partitioned into the training, validation, and a held-out validation set, with a ratio of 80% (n=1,030,438), 10% (n=129,127), and 10% (n=128,768). We constructed two cohorts to train the model: (1) a pretraining cohort for self-supervised learning for our foundation Transformer model; (2) an AD/ADRD/MCI finetuning cohort for training and evaluating the predictive model for AD/ADRD/MCI. The pretraining cohort includes all the patients in the training set (n=1,030,340). The AD/ADRD/MCI finetuning cohort only contains patients over the age 65 (by the end of records)(n=142,702). To simulate the predictive performance of the screening model in a retrospective manner, we used a sliding window for each patient to generate pairs of features and labels (see Figure 1e), where, given an index visit, the features were extracted from the EHR data prior to that index visit (feature window), and the labels were based on the EHR data following the index window (outcome window). Samples with no follow-ups by the end of the outcome window were considered censored and excluded. Patients who already had AD/ADRD/MCI by the end of the feature window were also excluded. In the analysis of AD/ADRD/MCI risk predictions in 5 years, we used a 1-year feature window and 5-year outcome window, which resulted in 445,142 records among 142,702 unique patients with 7.20% of those developing AD/ADRD/MCI in 5 years. These samples belonging to the training, validation, and held-out validation set of the full dataset formed the corresponding sets of AD/ADRD/MCI finetuning sets. Table 1 reports the demographic characteristics of both patient cohorts. The distributions of each patient’s length of presence, number of encounters, and medical codes are illustrated in Figure S1.

**Table 1.** Characteristics of the large-scale pretraining cohort and AD/ADRD/MCI finetuning cohort. The proportion or standard deviation is marked in parentheses.

| Characteristics | Pretraining Cohort | AD/ADRD/MCI<br>Finetuning Cohort |
| --- | --- | --- |
| Counts | 1,030,438 unique patients | 445,142 samples from<br>142,702 unique patients |
| Age (± SD) | 49.36 (± 24.48) | 73.44 (± 7.67) |
| Female | 607,513 (58.96) | 261,427 (58.72) |
| White | 568,954 (55.21) | 338,502 (76.04) |
| Black | 97,714 (9.48) | 32,371 (7.27) |
| Asian | 50,858 (4.94) | 15,103 (3.39) |
| Other Race | 106,029 (10.29) | 33,627 (7.55) |
| Unknown | 206,883 (20.07) | 25,539 (5.73) |
| Hypertension | 312,018 (30.28) | 232,519 (52.23) |
| Diabetes | 151,695 (14.72) | 93,649 (21.03) |
| Diabetes, complex | 12,482 (1.21) | 4,782 (1.07) |
| Hyperlipidemia | 335,805 (32.58) | 221,065 (49.66) |
| Any Vascular Risk | 490,224 (47.57) | 313,251 (70.37) |
| Statins | 241,655 (23.45) | 144,193 (32.39) |
| Aspirin | 184,348 (17.89) | 72,567 (16.30) |
| BP Meds | 299,616 (29.08) | 188,486 (42.34) |
| Any Vascular Meds | 408,258 (39.62) | 250,108 (56.18) |

### Identifying risk of AD/ADRD/MCI with EHR

We built a classification model on top of a Transformer network^19^, namely TRADE, to identify the risk of AD/ADRD/MCI based on the previous EHR (Figure 1d, see the Method section for more details). We assessed the predictive performance of our predictive models using two key metrics: the area under the receiver operating characteristic curve (AUC) and positive predictive values (PPV) at varying sensitivities (see Figure 2,4). For predicting the onset of AD/ADRD/MCI within 5 years from the reference index visit, the fine-tuned transformer achieved an AUROC of 0.735 (95% CI: 0.734-0.736) and average PPV (AP) of 0.199 (95% CI: 0.198-0.200). Since this predictive model aims to suggest cognitive screening for high-risk patients to providers, we also present PPVs of the high-risk patient groups predicted by the model (Figure 2). These high-risk groups are delineated by thresholds of the predicted risks from the top k% of patients with the highest predicted risks. For the 5-year prediction horizon, the PPVs are 39.2% and 27.8% and 22.4% for thresholds corresponding to the top 1, 5, and 10 percentiles, respectively. These values demonstrate the precision of the model at different screening operation points.

**Figure 2.**
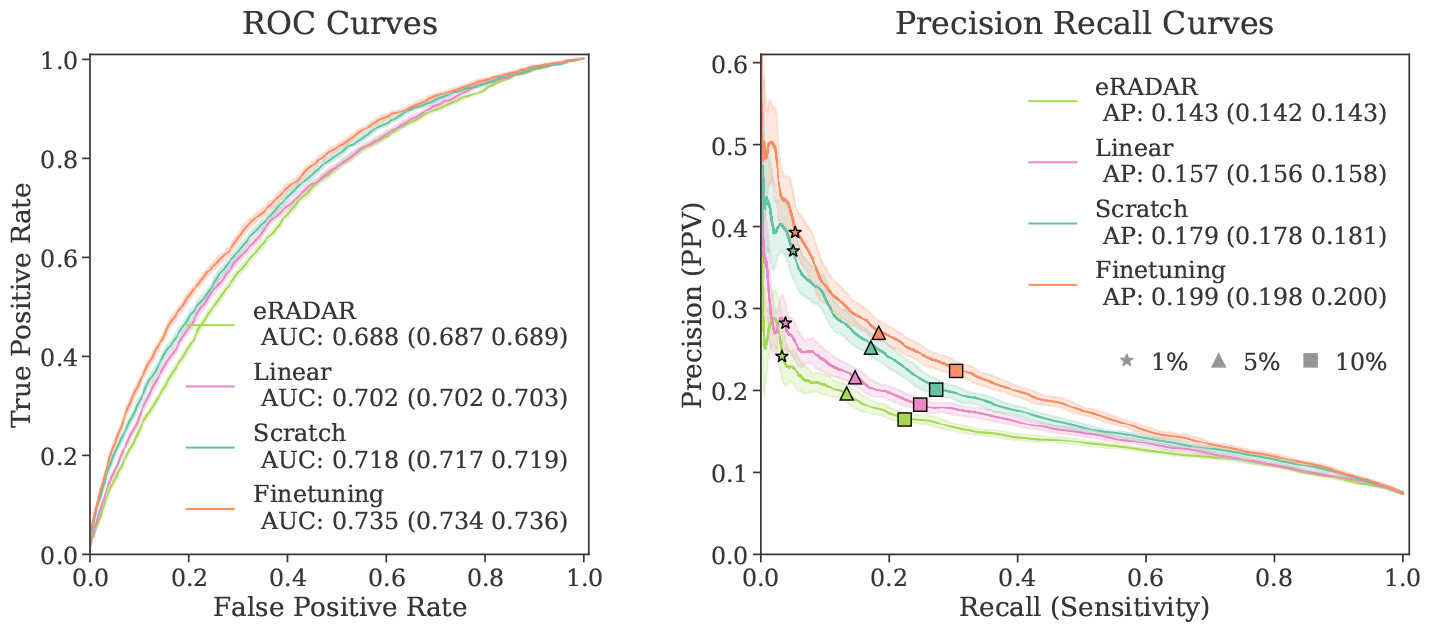
Performance on predicting AD/ADRD/MCI in 5 years with different methods: “Scratch” is training the Transformer from random initial weights; “Linear” is training a linear classifier on top of the foundation model representations; “Finetuning” is training the classifier layer together with the foundation model layers (see the Predictive modeling section for more detail). The performance is evaluated with ROC curves (left) and precision-recall curves (right). The PPVs and sensitivities of the patients with top 1, 5 and 10% risks are highlighted by ⋆, ▲ and ■, respectively.

Prior studies have also developed and validated EHR-based methods for early ADRD detection and prediction, yielding promising results. eRADAR^11^ is a statistical model estimating ADRD risks based on EHR variables, widely employed and validated in practice^13^. Comparing TRADE with eRADAR for predicting AD/ADRD/MCI over five years (see Figure 2), we observe that our foundation model significantly outperformed the statistical model which achieved AUROC of 0.688 (95% CI: 0.687-0.689) and AP of 0.143 (95% CI: 0.142, 0.143). Even without pre-training, our model achieves better performance, and linear probing (i.e only fine-tuning a linear classifier over the foundation model’s pre-learned features) bypasses the performance of the eRADAR model.

### Subcohort analysis

We compared the model performance of TRADE and eRADAR across various sub-cohorts according to gender, age, race, and other health conditions, like vascular risk (see Figure 3). Age and vascular risks are two major dementia risk factors significantly impacting prediction performance. Both models obtained higher PPVs among older patients and patients with vascular risks since AD/ADRD/MCIs were more prevalent among these subcohorts. However, the AUROCs were lower in the higher-risk subcohorts since it was harder to identify the negative patients among them. Notably, the AUROCs of eRADAR in age and vascular risk subgroups dropped more than those of TRADE. This phenomenon indicates that the predictions of eRADAR were mostly attributed to age and vascular diseases, while the Transformer model leveraged diverse medical information to further distinguish risks of patients of similar ages or health conditions.

**Figure 3.**
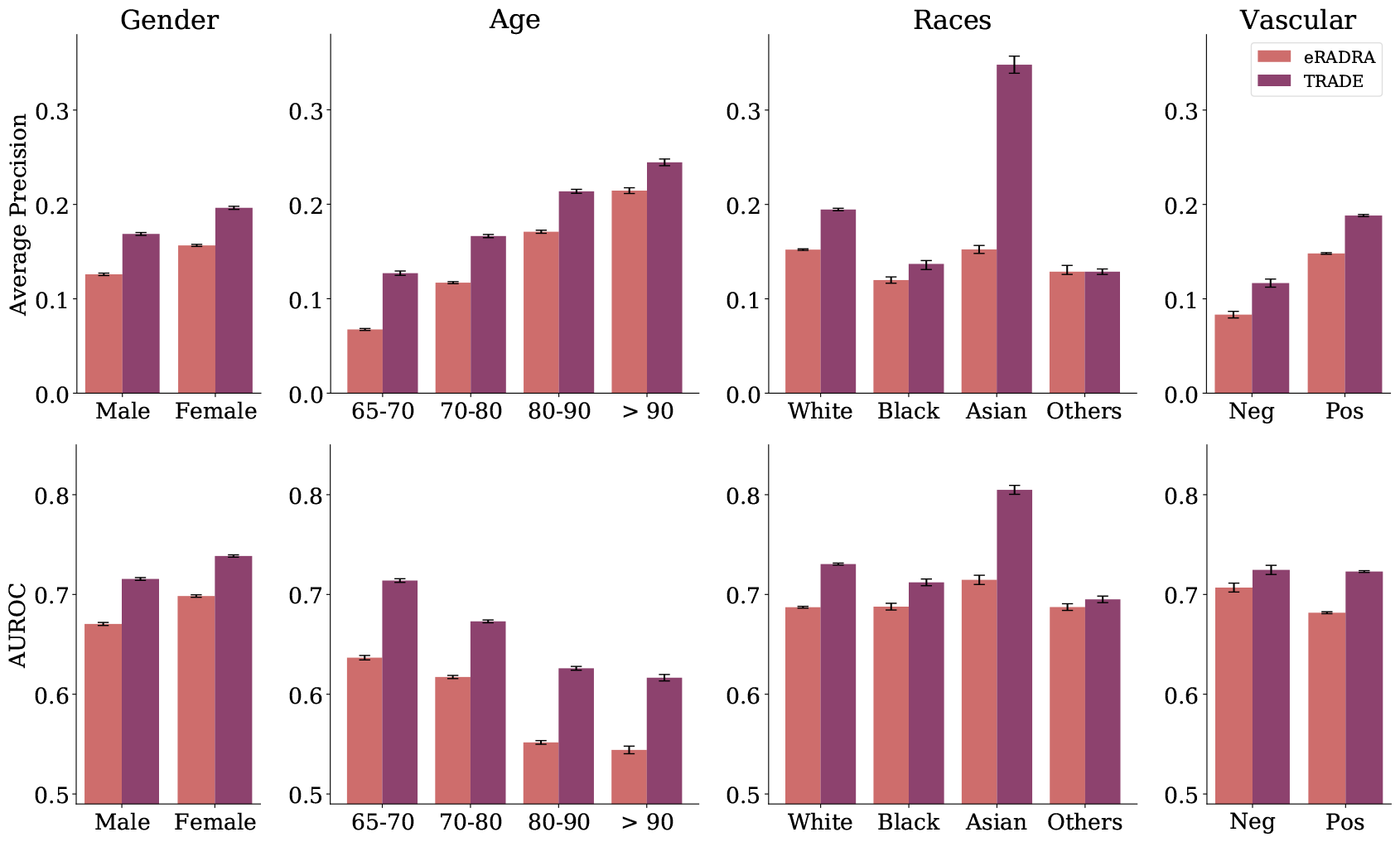
Performance across different subcohorts. Performance of the eRADAR (in orange) and of TRADE (in purple) for different subcohorts, partitioned by gender, age, race and vascular risk status. TRADE consistently outperformed eRADAR in every subcohort.

**Figure 4.**
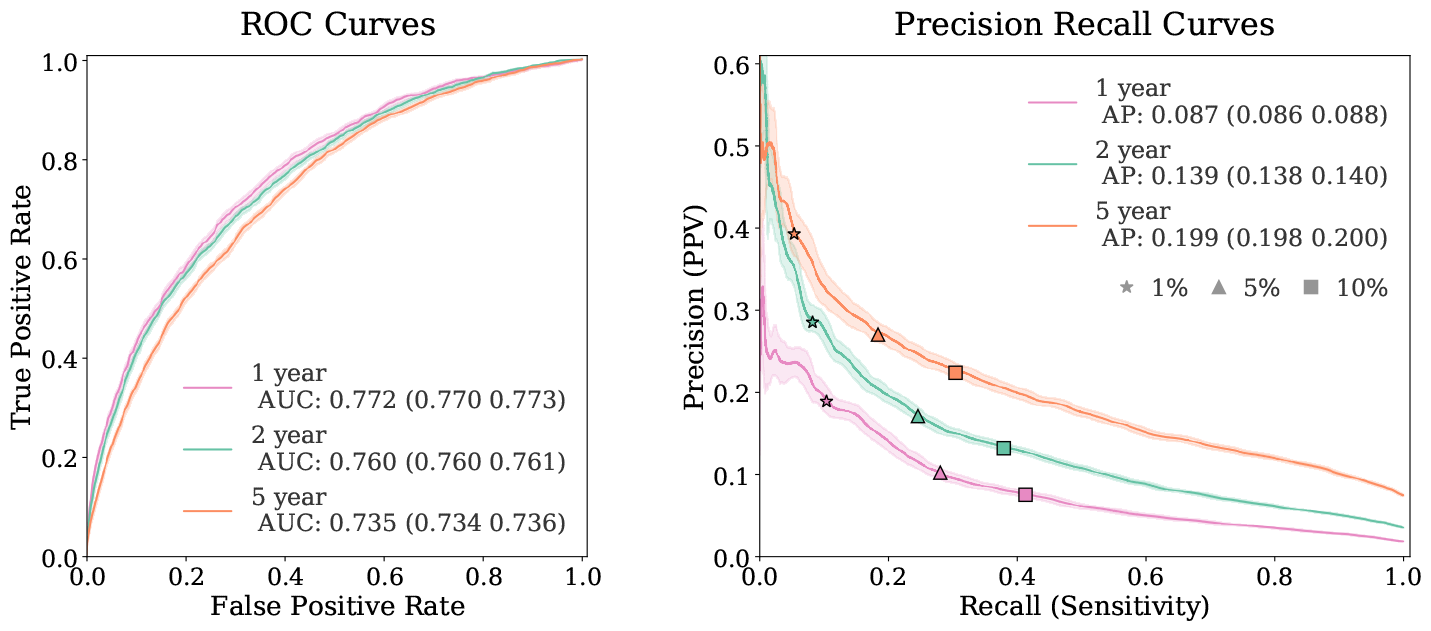
The comparison of TRADE performance on predicting AD/ADRD/MCI onset in different outcome windows. Prediction in the 1-year timeframe obtained the highest AUROC due to the least uncertainty. Prediction in the 5-year timeframe topped the PPVs as the corresponding prevalence was the highest. See Figure S2 for the comparison with baseline methods.

Among subcohorts separated by demographic characteristics, TRADE exhibited superior performance among females compared to males. Regarding the different races, while TRADE demonstrated lower performance among the underrepresented Black population when compared to the White population, it still consistently enhanced performance across all racial groups compared to eRADAR, with the most notable improvement observed among the Asian population. This finding suggests that TRADE has the potential to benefit all gender and race groups with a more accurate assessment of AD/ADRD/MCI risks.

### Improving performance with a pretrained foundation EHR model

A large-scale EHR dataset contains rich information on patients. To enable the model to understand EHR best, we designed a prediction framework with two stages (Figure 1a): (1) pretraining, where we pretrained a foundation model for EHR with Transformer architecture with the pretraining cohort. The model was trained without labels and merely by reconstructing randomly masked information from the EHR; (2) fine-tuning, where we fine-tuned the model with the medical history and AD/ADRD/MCI outcomes in the fine-tuning cohort to identify the high-risk patients (see the Method section for more details). We conducted pretraining only on the pretraining cohort. We fine-tuned the model with the training set in the AD/ADRD/MCI finetuning cohort and used the validation set to examine the performance for different hyperparameter settings, which guided model selection. The performance of the selected models was evaluated on the fully held-out validation set of patients and reported as an estimate of performance in new patient cohorts.

We compare the performance of the fine-tuned AD/ADRD/MCI predictive model with and without self-supervised pre-training. Figure 2 shows that under the same Transformer architecture, finetuning from pretrained foundation model obtained both higher AUROC and PPVs compared to training from scratch. In addition, a simplified linear classifier, trained over the representations extracted from the pre-trained foundation model, and comparable in complexity to eRADAR, still achieved superior performance to eRADAR. This demonstrates the value of a self-supervised pre-trained foundation AI model in building EHR-based predictive models.

### Prediction for different time intervals

The purpose of identifying patients at risk of dementia at different timelines varies. By predicting dementia across different time intervals, healthcare providers can intervene effectively at each stage. In this study, we developed models for predicting AD/ADRD/MCI onset within 1, 2, and 5 years from the visit being assessed. We constructed the samples of feature and label pair with different lengths of intervals for the outcome window (see Figure 1c). The corresponding characteristics of samples for various timelines are reported in Table S2. Figure 2 reports the prediction performance of the fine-tuned Transformer for different time intervals. Comparing the AUCs among them, prediction for the outcome window with 1 year tops with AUC at 0.772 (95% CI: 0.770, 0.773). This suggests the predictive model was more discriminative on the outcomes in the near future, while longer time intervals introduced more inherent uncertainty of the potential outcomes.

The precision-recall curves provide additional insights into the differences among outcome windows. Figure 2 demonstrates that the prediction on the longer outcome window had a high PPV compared to shorter ones at the same operation points. For patients at the top 1% risk level, the PPV of AD/ADRD/MCI onset in 5 years reached 39.2%, and decreased to 20.7% in the 1-year window. The false positive patients from the prediction in 1 year might develop dementia in the future and thus became the true positive in the 5-year window. On the other hand, the longer outcome window led to lower sensitivity as the early-stage AD/ADRD/MCI patients might not exhibit any risk factors at the current or past visits.

### Prediction with gap windows

Since the development and diagnosis of AD/ADRD/MCI is a prolonged process that can take several months, the recorded diagnosis on the EHR might be delayed. Considering the situation where the patient had onset of AD/ADRD/MCI, but has not yet been diagnosed, we conducted another analysis by excluding recently diagnosed patients within a gap window of 1 year after the end of the feature window, as shown in Figure 1e. Figure 5 shows the performance after the exclusion of these patients. As expected, the AUROC within 2 years dropped from 0.760 (95% CI: 0.760, 0.761) to 0.731 (95% CI: 0.730, 0.733), and the AUROC within 5 years dropped from 0.735 (95% CI: 0.734, 0.736) to 0.721 (95% CI: 0.720, 0.722). Less impact on the prediction in the 5-year time frame shows the ability of our model to identify the long-term AD/ADRD/MCI risks after eliminating the potential leakage due to the prolonged diagnosis process.

**Figure 5.**
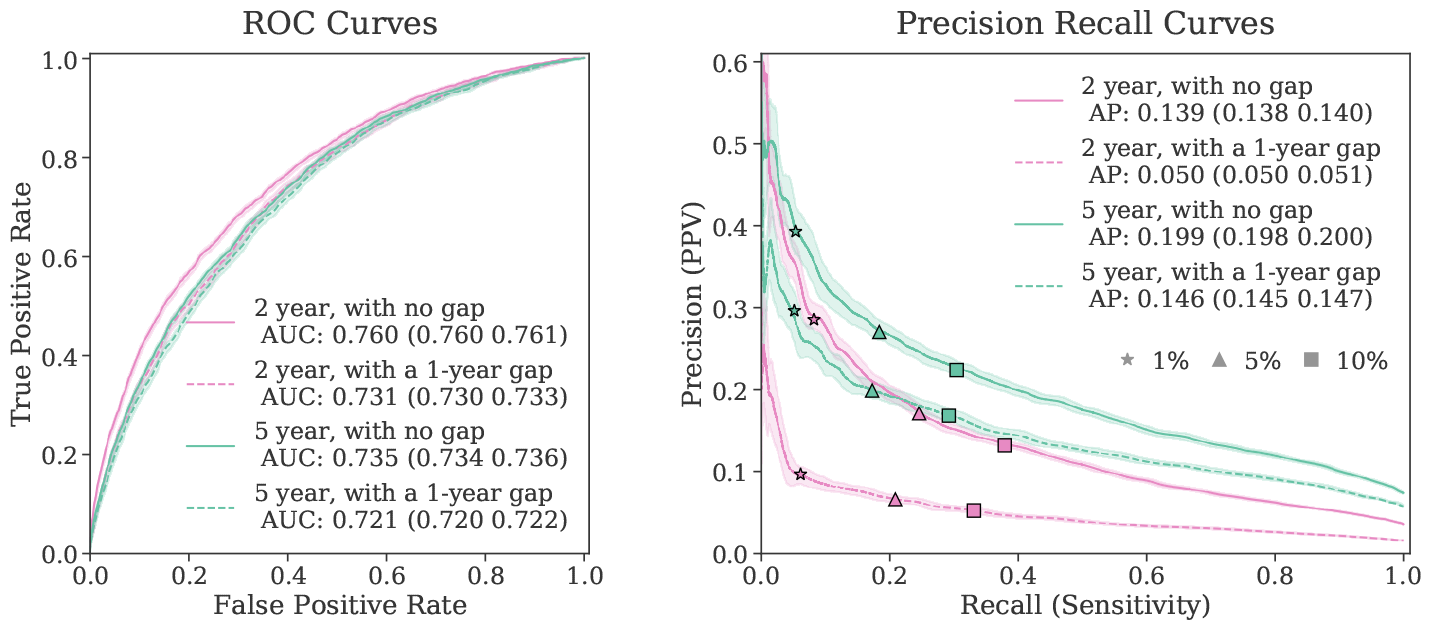
AD/ADRD/MCI prediction performance of TRADE, with and without excluding new-onset patients diagnosed with AD/ADRD/MCI within 1 year of the end of feature window. “1-year gap” indicates these samples were excluded, while “no gap” indicates no exclusion. The performance of the 5-year model dropped less than that of the 2-year model after the exclusion of onsets in the gap window.

### Model predictions correlate with cognitive impairment levels

We hypothesized that patients with higher predicted risk according to the model, if undergone cognitive screening, would also show a higher degree of impairment. From the technical standpoint, the correlation between the model score and cognitive test indicates the calibration of the model predictions and is a desired model behavior^28^. Figure 6 demonstrates the relationship between the risk estimated by the model and the Mini-Mental Status Exam (MMSE) cognitive scores^29^. For patients in the held-out validation set who had undergone MMSE examination in the 1-year time interval after the feature window, we extracted the MMSE scores mentioned in their clinical notes via ChatGPT with high accuracy^30^. We then compared these scores with the predicted risk from the 1-year AD/ADRD/MCI prediction Transformer model. As seen in Figure 6, the higher the risk model estimates, the more likely the patient would have severe cognitive impairment (i.e. MMSE < 20).

**Figure 6.**
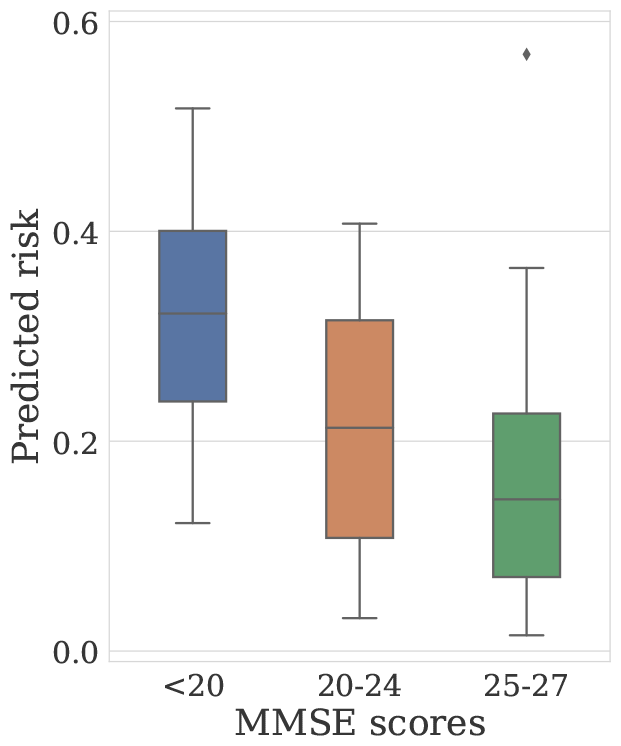
The analysis of Mini Mental State Exam (MMSE) cognitive scores in the heldout validation set, for patients undergone screening within 1 year from the index date. The box plot of predicted risk, according to the 1-year AD/ADRD/MCI prediction model, verse MMSE score ranges on patients with AD/ADRD/MCI outcomes, elucidates an association between higher predicted risks and lower MMSE scores.

### Model Interpretation

To understand the relationship between the model’s predictions and patients’ EHR tokens contributing to each predicted score, we computed the Integrated Gradients^31,32^of the prediction, with respect to the embedding layers of the EHR tokens. This provides a personalized explanation per patient. A higher magnitude of the integrated gradient for a variable indicates greater influence from that variable in the model output. These gradients for each patient can be aggregated to explain overall variables associated with a higher AD/ADRD/MCI risk. We note that this explanation method does not draw direct causal relationships. To aggregate the gradients, we use the mean of gradient norms across all positively labeled AD/ADRD/MCI patients in the heldout validation set as our metric. Tables 2 and S3 list the variables with the greatest magnitudes of gradients among the positively labeled AD/ADRD/MCI patients in the held-out validation set, unveiling the features most associated with high AD/ADRD/MCI risks. Only tokens with more than 10 occurrences were included to avoid outliers.

**Table 2.** The top 20 variables associated with future AD/ADRD/MCI diagnosis from 2- and 5-year models, with (gap=1), and without (gap=0) excluding patients whose AD/ADRD/MCI diagnosis occurred within 1 year of the end of the feature window. These ranked variables were extracted based on the magnitude of their gradients, and indicate an association rather than causation relationship between the variable and AD/ADRD/MCI risk. The ranking of variables is determined by the mean of gradient norms across all patients in the heldout validation set with a positive AD/ADRD/MCI label. The color of the cells indicates the category of variables based on either high-level ICD-10 categories or medication.

|  | AD/ADRD/MCI within 2 years | AD/ADRD/MCI within 2 years, with a 1-year gap | AD/ADRD/MCI within 5 years | AD/ADRD/MCI within 5 years, with a 1-year gap |
| --- | --- | --- | --- | --- |
| 1 | Other symptoms and signs involving cognitive functions and awareness R41.89 | Major depressive disorder, single episode, unspecified F32.9 | Parkinson's disease G20 | Major depressive disorder, single episode, unspecified F32.9 |
| 2 | Major depressive disorder, single episode, unspecified F32.9 | Personal history of malignant neoplasm of breast Z85.3 | Repeated falls R29.6 | Parkinson's disease G20 |
| 3 | Parkinson's disease G20 | Parkinson's disease G20 | Major depressive disorder F32.9 | Pure hypercholesterolemia E78.0 |
| 4 | Precordial pain R07.2 | Thrombocytopenia, unspecified D69.6 | Vascular parkinsonism G21.4 | Headache R51 |
| 5 | SERTRALINE | Pure hyperglyceridemia E78.1 | Urinary incontinence R32 | Vascular parkinsonism G21.4 |
| 6 | Abnormal weight loss R63.4 | Pure hypercholesterolemia E78.0 | ESCITALOPRAM OXALATE | Generalized anxiety disorder F41.1 |
| 7 | Dizziness and giddiness R42 | Estrogen receptor positive status [ER+] Z17.0 | LEVOTHYROXINE | Dizziness and giddiness R42 |
| 8 | Sick sinus syndrome I49.5 | Unspecified convulsions R56.9 | ATORVASTATIN | LEVOTHYROXINE |
| 9 | Unspecified urinary incontinence R32 | METOPROLOL SUCCINATE | LOSARTAN | Impaired glucose tolerance (oral) R73.02 |
| 10 | Urge incontinence N39.41 | Vascular parkinsonism G21.4 | Malignant neoplasm of prostate C61 | Mixed hyperlipidemia E78.2 |
| 11 | Syncope and collapse R55 | Overweight E66.3 | Personal history of malignant neoplasm of breast Z85.3 | Type 2 diabetes mellitus with unspecified complications E11.8 |
| 12 | Malignant neoplasm of prostate C61 | Dizziness and giddiness R42 | Dizziness and giddiness R42 | Precordial pain R07.2 |
| 13 | Peripheral vascular disease, unspecified I73.9 | Mixed hyperlipidemia E78.2 | Presence of automatic (implantable) cardiac defibrillator Z95.810 | Depression, unspecified F32.A |
| 14 | Mixed incontinence N39.46 | Type 2 diabetes mellitus with unspecified complications E11.8 | Cataract H26.9 | Acute posthemorrhagic anemia D62 |
| 15 | Transient cerebral ischemic attack, unspecified G45.9 | MELOXICAM | Nocturia R35.1 | Monoclonal gammopathy D47.2 |
| 16 | Hereditary and idiopathic neuropathy, unspecified G60.9 | Other general symptoms and signs R68.89 | Impaired glucose tolerance R73.02 | Vascular headache, not elsewhere classified G44.1 |
| 17 | Chronic kidney disease N18.4 | ATORVASTATIN | Symptoms involving the musculoskeletal system R29.898 | Anxiety disorder, unspecified F41.9 |
| 18 | Difficulty in walking R26.2 | Abnormal weight loss R63.4 | Presence of other heart-valve replacement Z95.4 | Hypo-osmolality and hyponatremia E87.1 |
| 19 | Vascular parkinsonism G21.4 | Impaired fasting glucose R73.01 | Obstructive sleep apnea G47.33 | Insomnia, unspecified G47.00 |
| 20 | Enlarged prostate with lower urinary tract symptoms N40.1 | Generalized anxiety disorder F41.1 | Type 2 diabetes mellitus with hyperglycemia E11.65 | Syncope and collapse R55 |

- D: Diseases of the blood and blood-forming organs - F: Mental, Behavioral and Neurodevelopmental disorders - I: Diseases of the circulatory system - R: Symptoms, signs and abnormal clinical and laboratory findings - E: Endocrine, nutritional and metabolic diseases - G: Diseases of the nervous system - N: Diseases of the genitourinary system - Medications

Several established and previously postulated risk factors of AD/ADRD/MCI emerge from these explanations, including vascular risk factors (such as heart disease, vascular disease, type 2 diabetes, hyperlipidemia, overweight)^33^, weight loss^34^, chronic kidney disease^35^, gait abnormality^36^, cataracts^37^, sleep apnea^38^, convulsions/seizures^39^, and mood disorder (including anxiety and depression)^40,41^. In addition to them, our model was able to recognize symptoms or diseases known to be precursors to AD/ADRD syndromes, including urinary incontinence, falls, and parkinsonism / Parkinson’s disease. Furthermore, certain medications were also highlighted, which may represent diagnoses that are AD/ADRD/MCI risk factors in the absence of formal diagnosis codes (e.g. escitalopram oxalate for depression or anxiety, atorvastatin for hyperlipidemia, metoprolol for hypertension), or may be linked via alternative relationships. This underscores the importance of utilizing interconnections among all EHR data to reveal a holistic picture of an individual’s health status.

## Discussion

Recent studies have consistently highlighted the delayed diagnosis of AD/ADRD in primary care^42^, often attributed to inadequate training and resource constraints. Notably, later diagnosis of AD/ADRD varies by socioeconomic, geolocation, racial and ethnic groups due to the disparities in access to timely diagnosis and specialists^43–45^. Previous research has revealed lower documentation rates of cognitive assessments 1-5 years prior to AD/ADRD diagnosis among Black/African American patients, older individuals, those with non-commercial health insurance, or residing in neighborhoods with lower mean income levels^46^. In this context, our study showcases the potential of TRADE, an EHR-based predictive model, to address this care gap by serving as a valuable tool for identifying high-risk patients who may benefit from targeted screening. Notably, our model exhibits improved performance compared to the best currently available tools, particularly among minority and high-risk groups, as illustrated in Figures 2 and 3.

While improvements were consistently observed across various demographic groups, disparities in accuracy persisted notably among racial subgroups, particularly between White and Black populations. While the difference in prevalence might partially explain the lower performance within the Black population, the higher precision among the less common Asian group contradicts such a hypothesis. This highlights the critical need not only for refining model development but also for recognizing the influence of social determinants of health^47,48^in deploying these models effectively. Our analysis delved into subcohort disparities in Mini-Mental Status Exam (MMSE) scores of patients who underwent screening and were diagnosed with AD/ADRD/MCI within the first year following the index date. In Figure 6a, we compared MMSE score distributions between White and Black subgroups, revealing a higher density in the Black group within the 20-24 MMSE range. While MMSE scores can be influenced by factors such as education level and education quality^49^, this finding may also suggest that a significant portion of Black patients may receive diagnoses at later disease stages. Indeed, the lower Positive Predictive Value (PPV) in the Black population may stem from delayed or underdiagnosis, further emphasizing the value of deploying decision support tools such as TRADE in the EHR systems in achieving a more inclusive screening program for all demographics.

In addition to promising performance for identifying patients at risk for AD/ADRD/MCI, our study includes an interpretability analysis, demonstrating that TRADE effectively extracts pertinent features from high-dimensional EHR data associated with an elevated AD/ADRD/MCI risk. The feature importance analysis underscores several established risk factors (e.g. vascular risks), and precursors strongly associated with AD/ADRD/MCI (e.g. monoclonal gammopathy). It is crucial to approach these associations with caution, recognizing them as correlations with potential AD/ADRD/MCI rather than direct causes. While further validation is needed to establish causality, these associations serve as valuable insights for driving future research into risk factors.

Our results demonstrate an improved performance of TRADE based on the self-supervised pre-trained foundation EHR model over the standard Transformer without pre-training. This suggests that our foundation EHR model has broader applicability in enhancing other EHR-based clinical tasks through fine-tuning with diverse datasets. Furthermore, while current EHR data predominantly includes generic diagnoses, lab values, and medications, integration of additional patient data such as medical images and clinical notes, if available, could offer richer contexts. The architecture of our foundation model allows seamless integration of multi-modal data. Building prediction frameworks capable of leveraging the extra information would be an important direction for future research.

In conclusion, we developed a risk assessment framework for AD/ADRD/MCI using structured EHR. Our findings demonstrate the efficacy of this model on the vast EHR of a healthcare system and its ability to significantly enhance the current clinical practices for the identification of AD/ADRD/MCI risks and screening programs. This improvement is particularly impactful for underrepresented and high-risk populations, where our model consistently outperforms existing methods. Meanwhile, we showed that the elevated predicted model scores are based on several recognizable risk factors or precursors of AD/ADRD/MCI. Finally, despite never directly training our model against cognitive screening scores such as MMSE, our results revealed that the higher risks predicted by the model do correlate with the degree of impairment, adding validity to the role of such models in the identification of undiagnosed AD/ADRD/MCI and tackling late diagnosis. Our approach to building predictive models based on the pre-trained foundation model of the EHR can benefit other diseases and similar risk assessment tasks, particularly in scenarios involving small and imbalanced patient cohorts.

## Methods

### Data

We used EHR data from NYU Langone Health including any visits (inpatient, outpatient and ED) between Jan 2013 and Jan 2023. This study has been approved by NYU Langone Institutional Review Board (IRB), as protocol s20-01095 Understanding and predicting Alzheimer’s Disease. Data was acquired from NYU Langone DataCore in a de-identified manner. Only patients with more than 5 visits were used in this study. This provided us the EHR of cohorts mentioned in the Study Participants section.

In the records of each patient, each visit was represented as a combination of variables, which include, demographic information (age, gender, race, ethnicity, 30 variables), alongside observed lab results (represented as LOINC codes, 86,529 variables total), medication orders (represented at Pharmaceutical class, 162,761 variables total) and diagnosis codes (represented as ICD-10 codes and SNOMED, 196,983 variables total). To convert these variables into tokens for Transformer inputs. We built the vocabulary list as follows: Each demographic category, diagnosis and medication code was treated as a unique token. The continuous lab values were binned into ranges of −10, −3, −1, −0.5, 0.5, 1, 3, 10 standard deviations from the population mean and then tokenized. Among all the variables, we selected 57,735 variables (6,482 lab values, 42,815 ICD-10 diagnosis codes, 8,387 medications, 50 demographics) with more than 100 occurrences. Any variables other than them were not included in the model. Moreover, we excluded ICD-10 codes related to amnesia, R41.1/2/3, to avoid potential leakage, which would not indicate AD/ADRD/MCI directly but could be used in preliminary diagnosis stages for these patients.

### Model development

#### ML architecture

As the state-of-the-art model for learning representations of the structured EHR data^21,22^, we adopted a Transformer^19^ network to encode the EHR at the patient level. We organized the longitudinal EHR data for each patient sequentially based on visit dates, resulting in a token sequence structured as follows (as illustrated in Figure 1c): “<CLS>“, {variables in 1st visit}, “<SEP>“, {variables in 2nd visit}, …, “<SEP>“. Here, “<CLS> and <SEP>” serve as the placeholder tokens representing the outputs used for classification and separating consecutive visits, respectively. Each medical observation within a visit was represented by a token, with each token encoded using a learnable embedding vector within the Transformer architecture. To capture the sequential nature and continuous variables across visits effectively, we introduced three learnable positional embeddings for each medical token, accounting for the patient’s age, the visit index, and the number of days since the beginning of the patient’s medical history. This refined approach offered a more nuanced representation of age and the temporal gap between encounters, compared to BERHT and Med-BERT^20,21^. Given that tokens within the same visit are permutation invariant, identical positional embeddings were applied to all tokens within a given visit. Then these embeddings were encoded by a Transformer neural network with 12 self-attention layers with 12 heads, 3072 dimensions for the feed-forward layers, and 786 dimensions for the encoder and pooling layers.

#### Model pretraining

Although the large-scale Transformer network has a strong capacity for encoding complicated high-dimensional data like EHR, it is also vulnerable to overfitting when trained on limited data for a single binary classification task, especially when the low prevalence in disease prediction tasks leads to limited positive samples^22^. To address this issue, we conducted self-supervised learning to pretrain the model with a large amount of unlabeled data. We employed the masked token modeling as proposed in Bidirectional Encoder Representations from Transformers (BERT)^24,25^for pretraining (Figure 1c) where the network was trained to predict randomly masked tokens (with a masking rate of 20%) from the model input. During pretraining, we used a sliding window framework to process each patient’s data. The first visit of the sliding window was randomly sampled, and we included the EHR data from that visit up to a future visit such that the number of tokens in the EHR segment would be fewer than 512. In each epoch, for each patient in the training set (n=1,030,438), one randomly sampled window was processed. The foundation model pre-training converged in 500 epochs. We used AdamW optimizer at batch size 256 and a learning rate 2 *×* 10^*−*5^ with a cosine learning rate decay. The computation was executed on 4 Nvidia A100 GPUs.

#### Predictive modeling for AD/ADRD/MCI

We trained the AD/ADRD/MCI predictive model, TRADE, based on the pretrained foundation Transformer. To conduct the retrospective analysis, we used a sliding window to create multiple samples from the medical trajectory of each patient (Figure 1e). We selected the index visits with at least 180 days stride and assumed them as the date for calculating the AD/ADRD/MCI prediction. The model took the EHR within 1 year before the index date as the model input. Multiple binary labels were created based on whether the patient was diagnosed with AD/ADRD/MCI within 1, 2, and 5 years after the index date. To avoid unrecorded diagnosis, we only kept samples that had follow-up encounters after the end of the outcome window. The max length of the tokens was also limited to 512 latest tokens in the input. The Transformer model was trained with the cross-entropy loss between the label and output of a linear classification layer at the Transformer encoding of the “<CLS>” token (see Figure 1d).

We use multiple ways to train the Transformer: (1) *scratch*, trained the Transformer with randomly initialized weights; (2) *linear probing* only updated the last linear layer of the Transformer while keeping the other layers frozen; (3) *Finetuning* used Low-Rank Adaptation (LoRA) method^26^ which injects and trains the rank decomposition matrices (with rank *r* = 128 and scale *α* = 256) into each linear layer of the Transformer architecture while keeping the original pretrained weights frozen. LoRA prevented the immediate overfitting and outperformed the vanilla finetuning method that updated all the parameters. We train the model for the downstream task for 10 epochs with an AdamW optimizer at batch size 64 and a learning rate 10^*−*5^ with a 10^*−*6^ weight decay. The computation was executed on 1 or 2 Nvidia V100 GPUs. These experiments were all early stopped by the optimal average PPVs on the validation set.

### eRADAR baselines

We established the benchmark for eRADAR prediction by evaluating it in the heldout validation set within AD/ADRD/MCI finetuning cohorts, using the same patients and index dates used for the Transformer model finetuning. We gathered the retrospective encounter data of variables used in the eRADAR model at the index dates for each individual, including age, sex, diagnoses in the past 2 years (e.g. congestive heart failure, cerebrovascular disease, diabetes (complex or any), etc.) based on ICD-10 codes, most recent vital signs for underweight (BMI <18.5), obese (BMI *≥*30), high blood pressure (*≥*140 SBP or*≥*90 DBP), healthcare utilization in the past 2 years (e.g. *≥*1 outpatient visit, *≥*1 emergency department visit, *≥*1 language and learning visit, etc), and utilization of medications in the past 2 years for non-tricyclic antidepressant and sedative-hypnotic. The eRADAR risks were computed with the scoring function and the published weights on each variable^11^.

### Extracting cognitive impairment level from clinical notes

Mini-Mental State Examination (MMSE)^29^ is an 11-question measure that tests the cognitive function of patients, where the scores range from 0-30 (28-30:Cognitively Normal, 25-27:MCI, 0-24:ADRD). We retrieved available MMSE scores of patients to study the association between estimated risks and cognitive impairment level. Among 44,222 samples from 14,131 patients identified as having AD/ADRD/MCI in the held-out validation set, 873 patients underwent MMSE cognitive test and were recorded by clinical notes at the institute of this study. From the clinical notes, we extracted the MMSE scores of the patients with OpenAI GPT-4^50,51^. A HIPAA-compliant private instance of ChatGPT was utilized to ensure data privacy.

### Performance metrics

We computed the area under the receiver operating characteristic (AUROC) and the positive predicted values (PPV) at different sensitivity levels, which are widely used for measuring the predictive accuracy of binary classification tasks. To report the statistical significance of descriptive statistics we computed 95% confidence intervals, and the bootstrapping method with 100 bootstrap iterations.

## Data availability

The trained TRADE model, its base foundation model, and corresponding code, notebooks can be made available upon direct request to the corresponding author.

## Data Availability

The original electronic health records will not be shared. The trained TRADE model, its base foundation model, and corresponding code, notebooks can be made available upon direct request to the corresponding author.

## Acknowledgements

W.Z., D.M., S.C., A.V.M. and N.R. were supported by the National Institute On Aging of the National Institutes of Health under Award R01AG079175. W.Z. received partial support from NSF Award 1922658. N.R., J.A.D., A.A.B, H.Z, A.V.M. was partially supported by the National Institute On Aging of the National Institutes of Health under Award R01AG085617.

N.R. and A.V.M are also supported by the National Institute On Aging of the National Institutes of Health under Award P30AG066512.

## Author contributions statement

N.R. led and supervised this study over all steps from design, development, and analysis. W.Z. and H.T. performed model development, training, validation, and analysis. A.V.M., J.A.D and A.A.B. provided clinical supervision throughout the study from design to analysis. S.C provided supervision on the deep learning model. H.Z. performed information retrieval from clinical notes. H.R.R, S.L.H, X.M, A.C., D.M., and E.A. performed analysis and preprocessing on EHR data. All authors participated in the writing of the manuscript.

## Additional information

The authors have declared no competing interests.

**Table S1.** Definition criteria for AD/ADRD/MCI onset.

| Condition | Criteria (ICD-10 codes and Medications) |
| --- | --- |
| AD/ADRD | F01.*: Any Vascular Dementia |
|  | F02.*: Dementia in other diseases classified elsewhere with or without behavioral disturbance |
|  | F03.*: Unspecified dementia with/without behavioral disturbance |
|  | F04.*: Amnestic disorder due to known physiological condition |
|  | G23.1: progressive supranuclear palsy |
|  | G30.*: Any Alzheimer's disease |
|  | G31.01: Pick's disease |
|  | G31.09: Other frontotemporal dementia |
|  | G31.83: Dementia with Lewy bodies |
|  | G31.9: Degenerative disease of nervous system, unspecified |
| Mild cognitive Impairment | G31.1: Senile degeneration of brain, not elsewhere classified |
|  | G31.84: Mild cognitive impairment of uncertain or unknown etiology |
|  | G31.85: Corticobasal degeneration |
| Dementia Medications | DONEPEZIL<br>GALANTAMINE<br>MEMANTINE<br>RIVASTIGMINE<br>TACRINE |

**Table S2.** The characteristics of AD/ADRD/MCI finetuning cohort for prediction in various timeframes.

| Characteristics | 1 year | 2 years | 5 years |
| --- | --- | --- | --- |
| Counts | 1,135,692 records from<br>223,653 unique patients | 1,000,433 records from<br>217,895 unique patients | 445,142 samples from<br>142,702 unique patients |
| Age (± SD) | 74.36 (± 7.29) | 74.13 (± 7.38) | 73.44 (± 7.67) |
| Female | 668,490 (58.86) | 588,000 (58.77) | 261,427 (58.72) |
| White | 845,843 (74.47) | 748,223 (74.79) | 338,502 (76.04) |
| Black | 87,701 (7.72) | 77,071 (7.70) | 32,371 (7.27) |
| Asian | 35,493 (3.13) | 31,667 (3.17) | 15,103 (3.39) |
| Other Race | 81,561 (7.18) | 72,289 (7.23) | 33,627 (7.55) |
| Unknown | 85,094 (7.49) | 71,183 (7.11) | 25,539 (5.73) |
| Hypertension | 630,256 (55.50) | 550,926 (55.07) | 232,519 (52.23) |
| Diabetes | 248,321 (21.87) | 217,892 (21.78) | 93,649 (21.03) |
| Diabetes, complex | 17,210 (1.52) | 14,657 (1.47) | 4,782 (1.07) |
| Hyperlipidemia | 605,379 (53.30) | 526,281 (52.61) | 221,065 (49.66) |
| Any Vascular Risk | 832,714 (73.32) | 728,977 (72.87) | 313,251 (70.37) |
| Statins | 398,058 (35.05) | 350,195 (35.00) | 144,193 (32.39) |
| Aspirin | 166,253 (14.64) | 152,474 (15.24) | 72,567 (16.30) |
| BP Meds | 495,889 (43.67) | 439,045 (43.89) | 188,486 (42.34) |
| Any Vascular Meds | 656,821 (57.83) | 580,701 (58.05) | 250,108 (56.18) |

**Table S3.** The top 20 variables associated with future AD/ADRD/MCI diagnosis within 5 years. Each column reports variables across patients with varied time-to-dementia from the index date. The color of the cells indicates the category of variables based on either high-level ICD-10 categories or medication.

|  | AD/ADRD/MCI within 0-1 year | AD/ADRD/MCI within 1-2 years | AD/ADRD/MCI within 2-3 years | AD/ADRD/MCI within 3-5 years |
| --- | --- | --- | --- | --- |
| 1 | Altered mental status, unspecified R41.82 | Precordial pain R07.2 | Precordial pain R07.2 | SERTRALINE |
| 2 | Precordial pain R07.2 | Parkinson's disease G20 | Parkinson's disease G20 | Abnormal weight loss R63.4 |
| 3 | Epilepsy, unspecified, not intractable, without status epilepticus G40.909 | Abnormal weight loss R63.4 | Vascular parkinsonism G21.4 | Unspecified urinary incontinence R32 |
| 4 | Major depressive disorder, single episode, unspecified F32.9 | Vascular parkinsonism G21.4 | Unspecified convulsions R56.9 | Parkinson's disease G20 |
| 5 | Parkinson's disease G20 | LOSARTAN | Presence of automatic (implantable) cardiac defibrillator Z95.810 | Repeated falls R29.6 |
| 6 | Vascular parkinsonism G21.4 | Major depressive disorder, single episode, unspecified F32.9 | Abnormal weight loss R63.4 | Vascular parkinsonism G21.4 |
| 7 | Unspecified urinary incontinence R32 | Nocturia R35.1 | Major depressive disorder, single episode, unspecified F32.9 | Major depressive disorder, single episode, unspecified F32.9 |
| 8 | INR in Platelet poor plasma by Coagulation assay [+3-10 sd] | DIGOXIN | Unspecified urinary incontinence R32 | Presence of automatic cardiac defibrillator Z95.810 |
| 9 | Nocturia R35.1 | ATORVASTATIN | Syncope and collapse R55 | Unspecified cataract H26.9 |
| 10 | Hereditary and idiopathic neuropathy, unspecified G60.9 | Dizziness and giddiness R42 | Hereditary and idiopathic neuropathy, unspecified G60.9 | Other secondary pulmonary hypertension I27.2 |
| 11 | ESCITALOPRAM OXALATE | Unspecified urinary incontinence R32 | LOSARTAN | MECLIZINE |
| 12 | Dizziness and giddiness R42 | Malignant neoplasm of prostate C61 | DIGOXIN | Syncope and collapse R55 |
| 13 | Urge incontinence N39.41 | Functional dyspepsia K30 | Dizziness and giddiness R42 | Dizziness and giddiness R42 |
| 14 | End stage renal disease N18.6 | Hereditary and idiopathic neuropathy, unspecified G60.9 | Difficulty in walking, not elsewhere classified R26.2 | Hereditary and idiopathic neuropathy, unspecified G60.9 |
| 15 | Abnormal weight loss R63.4 | ATORVASTATIN | Nocturia R35.1 | Headache R51 |
| 16 | CARBIDOPA-LEVODOPA | LEVOTHYROXINE | Overactive bladder N32.81 | Hematuria, unspecified R31.9 |
| 17 | Unspecified cataract H26.9 | Headache R51 | Unspecified cataract H26.9 | DIGOXIN |
| 18 | LOSARTAN | CLONAZEPAM | Unspecified atherosclerosis of native arteries of extremities, other extremity I70.208 | Nocturia R35.1 |
| 19 | Hematuria, unspecified R31.9 | CARBIDOPA-LEVODOPA | CARVEDILOL | FUROSEMIDE |
| 20 | Syncope and collapse R55 | Hematuria, unspecified R31.9 | Obstructive sleep apnea G47.33 | Polymyalgia rheumatica M35.3 |

- D: Diseases of the blood and blood-forming organs - E: Endocrine, nutritional and metabolic diseases - F: Mental, Behavioral and Neurodevelopmental disorders - G: Diseases of the nervous system - I: Diseases of the circulatory system - N: Diseases of the genitourinary system - R: Symptoms, signs and abnormal clinical and laboratory findings - Medications

**Figure S1.**
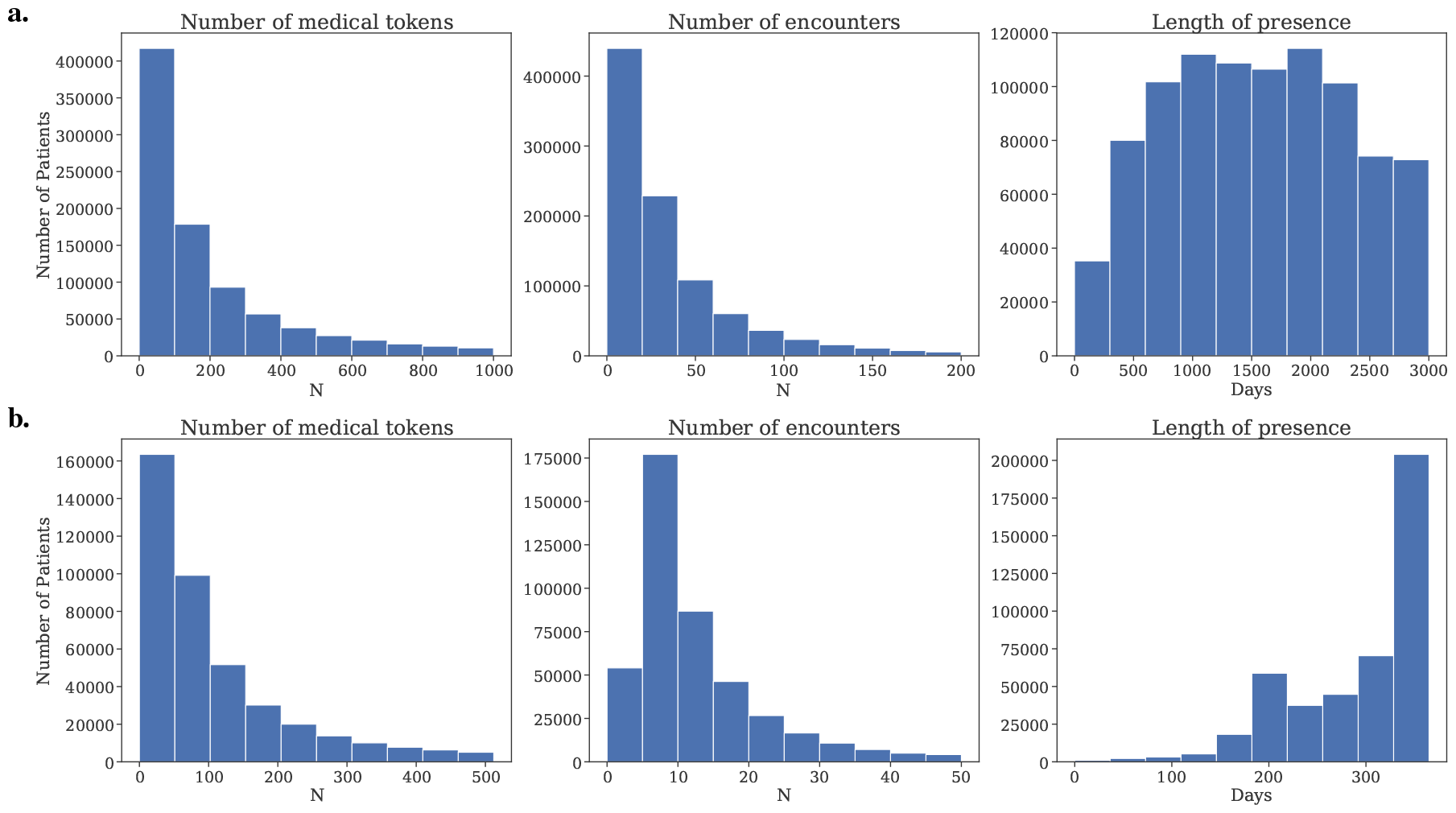
The histograms (from left to right) show the distributions of the number of medical tokens, the number of encounters, and the length of presence, respectively, for: **a**. full EHR data of each patient in the pretraining cohort (N=1,030,438 patients) **b**. EHR data in the *1-year feature window* of the (N=445,142) samples from 142,702 unique patients used during the AD/ADRD/MCI finetuning.

**Figure S2.**
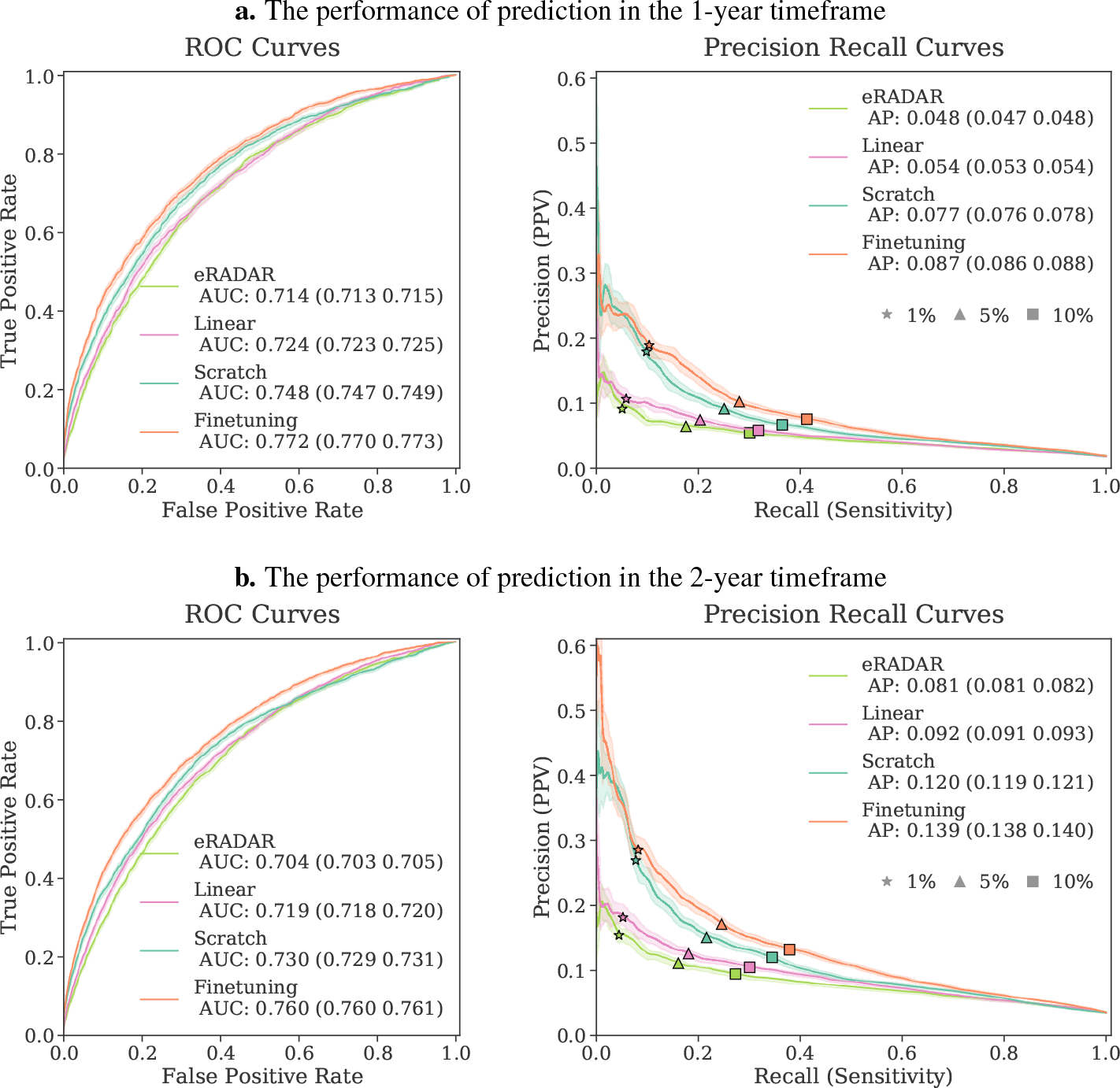
The comparison of performance on predicting AD/ADRD/MCI onset in a. 1-year and b. 2-year outcome windows with different baselines. The finetuned foundation model consistently outperformed the other baselines.

**Figure S3.**
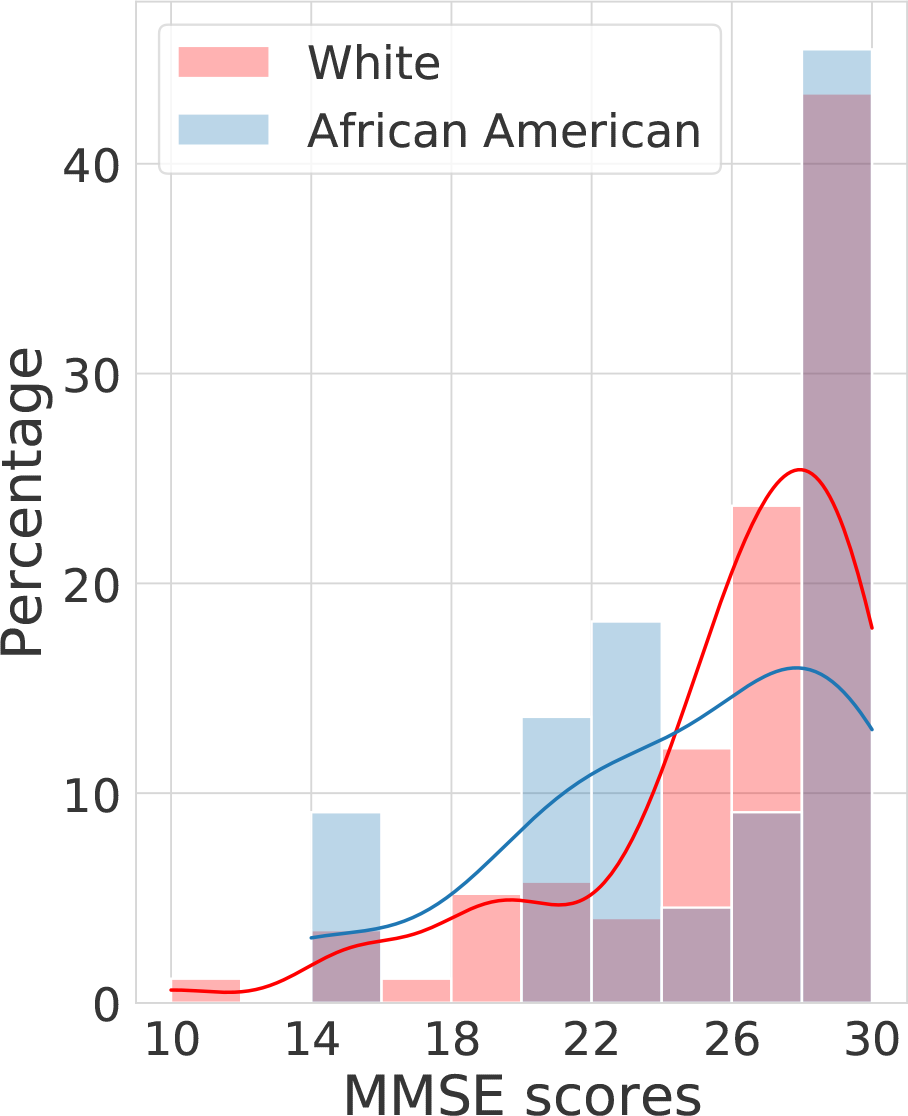
The analysis of Mini Mental State Exam (MMSE) cognitive scores in the heldout validation set, for patients who underwent screening and were diagnosed with AD/ADRD/MCI within 1 year from the index date. The distribution of scores for AD/ADRD/MCI patients in White vs Black implies that Black patients had lower MMSE scores at diagnosis time compared to white patients, potentially revealing delayed diagnosis in these populations.

